# Primary Care Diagnosis and Treatment of Attention-Deficit/Hyperactivity Disorder in School-aged Children: Trends and Disparities During the COVID-19 Pandemic

**DOI:** 10.1101/2021.11.03.21265902

**Authors:** Yair Bannett, Alex Dahlen, Lynne C. Huffman, Heidi M. Feldman

## Abstract

**Importance:** Little is known about changes in health care in the first year of the pandemic for the large population of school-aged children with attention-deficit/hyperactivity disorder (ADHD), who were especially impacted by lockdowns, school closures, and remote learning.

**Objective:** To assess temporal trends in rates of primary care provider (PCP) diagnosis and treatment of school-aged children with ADHD in the first year of the COVID-19 pandemic as compared to pre-pandemic years, and to investigate disparities in care.

**Method:** We retrospectively analyzed electronic health records from all primary care visits (in-person and telehealth) of children ages 6-17 years seen between 01/2016 and 03/2021 in a community-based primary healthcare network in California (n=77,298 patients). Study Outcomes: (1) # of primary care visits, (2) # of visits with ADHD diagnosis (ADHD-related visits), (3) # of first ADHD diagnoses, (4) # of PCP prescriptions for ADHD medications (stimulants, alpha-2 agonists, atomoxetine), (5) # of first PCP prescriptions of ADHD medications. Interrupted time-series analysis evaluated changes in rates of study outcomes during 4 quarters of the pandemic year (3/15/2020-3/15/2021) compared to pre-pandemic years. Patient demographic characteristics were compared pre-pandemic to pandemic year.

**Results:** In the first quarter (Q1) of the pandemic year, all primary care visits dropped by 62% (CI 54.9-67.2%); ADHD-related visits dropped by 33% (95% CI 22.2-43.6%). In Q2-4, while all primary care visits remained significantly below pre-pandemic rates, ADHD-related visits returned to pre-pandemic rates. Conversely, rates of first ADHD diagnoses remained at half of pre-pandemic rates throughout the year (Q1-4). ADHD medication prescription rates remained stable throughout the pandemic year. The proportion of patients living in low-income neighborhoods who received ADHD-related care (ADHD-related visits and first ADHD diagnoses) were lower during the pandemic year compared to pre-pandemic years. Females comprised a higher proportion of first ADHD diagnoses compared to pre-pandemic years (34% vs. 28%, absolute standardized difference=0.13, p=0.03).

**Conclusion:** Ongoing treatment for school-aged children with ADHD was maintained during the pandemic, especially in children from high-income families. Socioeconomic differences in ADHD-related care emphasize the need to improve access to care for all children with ADHD in the ongoing pandemic and beyond.

## Introduction

School-aged children with attention-deficit/hyperactivity disorder (ADHD) were particularly impacted by lockdowns, school closures, and remote learning in the first year of the COVID-19 pandemic.^1-3^ Most US children with ADHD are diagnosed and treated by their primary care provider (PCP).^4^ However, little is known about the pandemic’s effect on the diagnosis and treatment of school-aged children with ADHD by PCPs. Examining trends and investigating disparities in ADHD diagnosis and treatment during the pandemic is important because it can inform prioritization of ADHD-related care and identify intervention targets to reduce disparities during the ongoing pandemic.

We aimed to assess rate of PCP diagnosis and treatment of school-aged children with ADHD during the pandemic as compared to pre-pandemic years and to investigate disparities in ADHD-related care. We hypothesized that, compared to pre-pandemic years, ADHD-related care utilization will be high throughout the first year of the pandemic due to the increased impact on children with ADHD. We further hypothesized that first-time ADHD diagnoses will increase in the second half of the pandemic year, as lockdowns were lifted and schools gradually reopened.

## Methods

We retrospectively analyzed electronic health record (EHR) data from Packard Children’s Health Alliance (PCHA), a community-based pediatric healthcare network in the San Francisco Bay Area, affiliated with Stanford Children’s Health and Lucile Packard Children’s Hospital, serving ∼100,000 children annually, previously described.^5^ Structured EHR data were extracted through the STAnford Medicine Research data Repository Observational Medical Outcomes Partnership (OMOP) Common Data Model (CDM) [STARR-OMOP].^6^ The Stanford University School of Medicine institutional review board approved the study.

We included data from all primary care visits (in-person and telehealth) of children ages 6-17 years seen for at least 6 months between 01/2016 and 03/2021 with a visit diagnosis of ADHD (n=77,298 patients).

### Study outcomes

We examined temporal trends in five outcomes: (1) # of all primary care visits, (2) # of visits with ADHD diagnosis (“ADHD-related visits”), (3) # of first ADHD diagnoses, (4) # of PCP prescriptions for ADHD medications (stimulants, alpha-2 agonists, atomoxetine), and (5) # of first PCP prescriptions for ADHD medications.

ADHD-related visits and prescriptions for ADHD medications were identified based on OMOP CDM concepts. ADHD-related visits were identified by the OMOP code for ADHD (concept id = 43409) or one of its decendent concepts, except that diagnoses of hyperkinesis (438132, 437261, and descendants) were excluded. For each patient, a first ADHD diagnosis was defined as the first ADHD-related visit in the study period (01/2016-03/2021).

ADHD medication was identified by OMOP code for methylphenidate (705944), guanfacine (1344965), dextroamphetamine (719311), dexmethylphenidate (731533), clonidine (1398937), atomoxetine (742185), amphetamine (714785), or any descendant concept.

### Independent Variables

We used patient structured data in the EHR to describe patient age (mean age in years during study period), sex, race (White/Asian/African American/Other), ethnicity (Hispanic/Non-Hispanic), and medical insurance type at first study period visit (private/public). Median household income was calculated from the patient’s ZIP code based on 2019 census data,^7^ bucketed into quartiles.

### Statistical analysis

Interrupted time-series analysis evaluated changes in rates of study outcomes. The causal effect of the pandemic year was parameterized in four step-function changes at regular 3-month intervals after lockdown: 3/15/20, 6/15/20, 9/15/20, and 12/15/20. For each metric, we used an interrupted time series approach on the log of the metric in order to determine % change between the actual data and the counterfactual data that likely would have happened without the pandemic. We used yearly differencing to control for seasonal patterns in the data, and the logarithm resulted in residuals that were approximately normal. The model was an ordinary least squares with eight terms: a linear term, three quarterly nuisance parameters (Jan-Mar, Apr-Jun, and Jul-Oct), as well as the 4 indicator variables for each quarter of the pandemic year. Confidence intervals were autocorrelation robust. In addition to this primary model, we we conducted a sensitivity analysis that looked at a simpler model (no nuisance parameters) and a more complex model (seasonal ARIMA); both gave similar results.

Univariate analyses compared patient demographic characteristics pre-pandemic and pandemic, using t-test for continuous variables and chi-square for categorical variables. All analyses were conducted using Python, version 3.8.5.

## Results

Of 77,298 eligible children, 4,808 (6.2%) had an ADHD diagnosis anytime in the study period with an average age at diagnosis of 10.7 years (SD 3.4 years). Of those, 3,443 (72%) were male, 1,779 (37%) non-hispanic white, 2,786 (58%) privately insured, and 2,574 (54%) were prescribed an ADHD medication (**Table 1**).

**Table 1.** Characteristics of patient cohort aged 6-17 years (n=77,298)

| n | No ADHD<br>72,490 | ADHD<br>4,808 | Absolute<br>Standardized<br>Difference<sup>a</sup> |
| --- | --- | --- | --- |
| Age<sup>b</sup> | 13.9 (4.2) | 15.1 (3.9) | 0.28 |
| Male, n (%) | 35,520 (49.0%) | 3,443 (71.6%) | 0.45 |
| Race / Ethnicity |  |  |  |
| non-Hispanic White | 18,806 (25.9%) | 1,779 (37.0%) | 0.25 |
| non-Hispanic Black | 2,198 (3.0%) | 211 (4.4%) | 0.08 |
| Hispanic | 7,925 (10.9%) | 586 (12.2%) | 0.04 |
| Asian | 10,757 (14.8%) | 339 (7.1%) | 0.22 |
| Other<sup>c</sup> | 10,430 (14.4%) | 583 (12.1%) | 0.06 |
| Unknown | 22,374 (30.9%) | 1,310 (27.2%) | 0.08 |
| Insurance Type, n (%) |  |  |  |
| Public | 26,245 (36.2%) | 1,786 (37.1%) | 0.02 |
| Private | 41,104 (56.7%) | 2,786 (57.9%) | 0.03 |
| Unknown | 5,141 (7.1%) | 236 (4.9%) | 0.09 |
| Household Income Quartile<sup>d</sup> |  |  |  |
| Bottom quartile (< \$99,955) | 17,993 (24.8%) | 1,281 (26.6%) | 0.04 |
| Second quartile (>99,955-\$133,381) | 17,620 (24.3%) | 1,269 (26.3%) | 0.05 |
| Third quartile (\$133,381- \$153,429) | 18,121 (24.9%) | 1,077 (22.4%) | 0.06 |
| Top quartile (> \$153,429) | 17,781 (24.5%) | 1,086 (23.0%) | 0.05 |
| No income data available | 975 (1.3%) | 95 (2.0%) | - |

<sup>a</sup> Absolute Standardized Difference values of 0.2, 0.5, and 0.8 correspond to small, moderate, and large differences, respectively.
<sup>b</sup> Age in years on January 1, 2020. Mean and standard deviation is displayed for continuous variables.
<sup>c</sup> Includes Other, Native American and Pacific Islander.
<sup>d</sup> Calculated from the patient's ZIP code based on 2019 census data bucketed into quartiles.

In the first quarter (Q1) of the pandemic year, all primary care visits dropped by 62% (CI 54.9-67.2%); ADHD-related visits dropped by 33% (95% CI 22.2-43.6%). In Q2-4, while all primary care visits remained significantly below pre-pandemic rates, ADHD-related visits returned to pre-pandemic rates (**Figure**). Conversely, rates of first ADHD diagnoses remained at half of pre-pandemic rates throughout the year (Q1-4).

**Figure 1.**
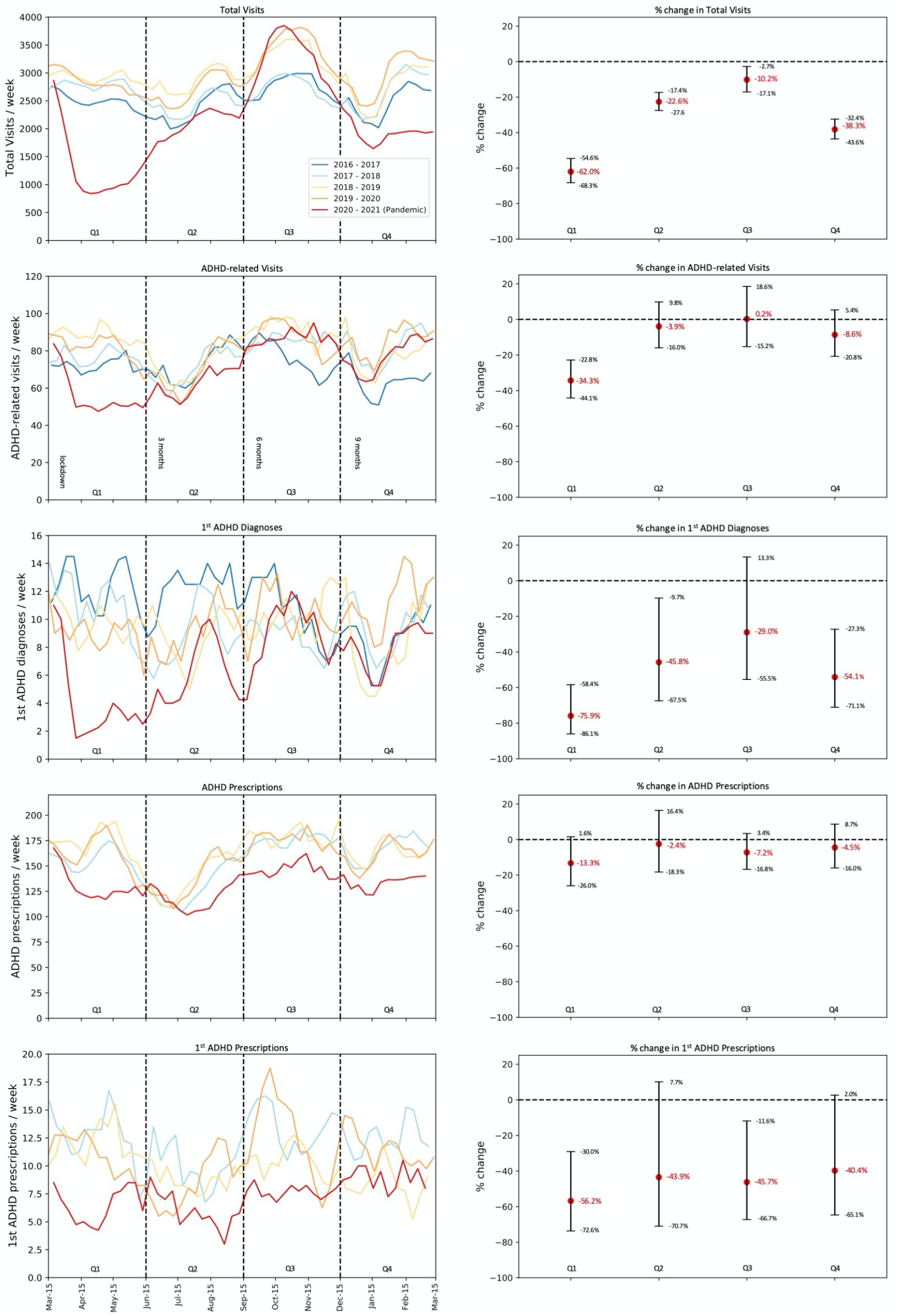
Multi-year temporal trends in all primary care visits and the four study outcomes (left) and corresponding results of Interrupted Time series Analysis (right).

Prescription rates of ADHD medications dropped minimally in the pandemic year. However, first prescription of ADHD medications remained significantly below pre-pandemic rates throughout the year (**Figure**).

As shown in **Table 2**, the proportion of patients living in low-income neighborhoods who received ADHD-related care (ADHD-related visits and first ADHD diagnoses) were lower during the pandemic year compared to pre-pandemic years (22% vs. 27% for ADHD-related visits, Absolute Standardized Difference (ASD)=0.12; 23% vs. 31% for first ADHD diagnoses, ASD=0.16, p<0.001). Females comprised a higher proportion of first-time ADHD diagnoses compared to pre-pandemic years (34% vs. 28%, ASD=0.13, p=0.03). Differences in race/ethnicity and insurance type were small. In post-hoc interrupted time-series analysis stratified by income quartile (data not shown), patients from the highest income quartile returned to pre-pandemic rates of ADHD-related care in Q2-4; those from the lowest quartile experienced continued disruption in care.

**Table 2.** Comparing patient demographics before and after the pandemic for number of ADHD-related visits and for first ADHD diagnoses.^a^

|  | ADHD-related Visits |  |  |  | 1<sup>st</sup> ADHD diagnoses |  |  |  |
| --- | --- | --- | --- | --- | --- | --- | --- | --- |
|  | Pre-pandemic<br>(4 years) | Pandemic<br>(1 year) | p | ASD | Pre-pandemic<br>(4 years) | Pandemic<br>(1 year) | p | ASD |
| **n** | 17,147 visits | 3,643 visits |  |  | 2,079 patients | 338 patients |  |  |
| **Age at visit/diagnosis, avg (std)** | 11.9 (3.1) | 12.0 (3.0) | 0.18 | 0.02 | 10.9 (3.3) | 11.1 (3.2) | 0.40 | 0.05 |
| **Female, n (%)** | 4,255 (24.8%) | 976 (26.8%) | 0.01 | 0.05 | 577 (27.8%) | 114 (33.7%) | 0.03 | 0.13 |
| **Race / Ethnicity, n (%)** |  |  | < 0.0001 |  |  |  | 0.03 |  |
| non-Hispanic White | 6,719 (39.2%) | 1,353 (37.1%) |  | -0.04 | 743 (35.7%) | 109 (32.2%) |  | -0.07 |
| non-Hispanic Black | 697 (4.1%) | 101 (2.8%) |  | -0.07 | 64 (3.1%) | 4 (1.2%) |  | -0.11 |
| Hispanic | 2,209 (12.9%) | 452 (12.4%) |  | -0.01 | 242 (11.6%) | 33 (9.8%) |  | -0.06 |
| Asian | 964 (5.6%) | 245 (6.7%) |  | 0.05 | 125 (6.0%) | 30 (8.9%) |  | 0.12 |
| Other | 2,160 (12.6%) | 419 (11.5%) |  | -0.03 | 236 (11.4%) | 35 (10.4%) |  | -0.03 |
| Did Not Specify / Unknown | 4,398 (25.6%) | 1,073 (29.5%) |  | 0.09 | 669 (32.2%) | 127 (37.6%) |  | 0.11 |
| **Insurance Type, n (%)** |  |  | 0.02 |  |  |  | 0.79 |  |
| Public | 6,538 (38.1%) | 1,348 (37.0%) |  | -0.02 | 767 (36.9%) | 129 (38.2%) |  | 0.03 |
| Private | 10,148 (59.2%) | 2,169 (59.5%) |  | 0.01 | 1,204 (57.9%) | 194 (57.4%) |  | -0.01 |
| Unknown | 461 (2.7%) | 126 (3.5%) |  | 0.05 | 108 (5.2%) | 15 (4.4%) |  | -0.03 |
| **HH Income Quartile, n (%)** |  |  | < 0.0001 |  |  |  | 0.04 |  |
| bottom (< \$99,955) | 4,624 (27.0%) | 785 (21.5%) | | -0.12 | 641 (30.8%) | 79 (23.4%) | | -0.16 |
| second (\$99,955 - \$133,381) | 4,975 (29.0%) | 965 (26.5%) | | -0.06 | 508 (24.4%) | 90 (26.6%) | | 0.05 |
| third (\$133,381 - \$153,429) | 3,786 (22.1%) | 984 (27.0%) | | 0.12 | 461 (22.2%) | 80 (23.3%) | | 0.04 |
| top (> \$153,429) | 3,762 (21.9%) | 909 (25.0%) | | 0.07 | 469 (22.6%) | 89 (26.3%) | | 0.09 |

<sup>a</sup> Comparisons are made using a two-sided t-test for continuous variables, and using a chi-square test for categorical ones.
<sup>b</sup> ASD=Absolute Standardized Difference; values of 0.2, 0.5, and 0.8 correspond to small, moderate, and large differences, respectively.
<sup>c</sup> Age in years for all ADHD-related visits and for first ADHD diagnosis. Mean and standard deviation is displayed for continuous variables.

## Discussion

Throughout the first year of the pandemic, overall ADHD-related care for school-aged children was relatively maintained. The findings suggest that families perceived follow-up and medication treatment for children with ADHD as necessary. However, children from low-income families experienced prolonged disruption in ADHD-related care during the pandemic, suggesting worsening of previously reported disparities in ADHD care.^8^ Low rates of first ADHD diagnoses throughout the pandemic may relate to decreased recognition of ADHD symptoms or increased uncertainty about diagnosis in remote learning settings. Increased proportion of females with first ADHD diagnoses may relate to lower rates of disruptive behaviors in males – that often prompt evaluations by PCPs – or increased recognition in females during remote learning.

Study limitations include reliance on EHR data from primary care settings, which did not include sub-specialist care. Our dataset lacked an indicator for telehealth visits, precluding the assessment of telehealth access as a contributor to care disparities.

## Conclusion

Study findings shed light on changes in health care provided to children with ADHD and their families. Socioeconomic differences in ADHD-related care emphasize the need to improve access to care for all children with ADHD in the ongoing pandemic and beyond.

## Data Availability

Datasets used for this study must be completely de-identified according to federal regulations prior to release for sharing. Given the potentially sensitive nature of these data, we will make the datasets available only under a data-sharing plan. Requests to use the information and data from this study will be considered on a case-by-case basis, following written request to the corresponding author.

## Acknowledgments

This research used data or services provided by STARR, “STAnford medicine Research data Repository,” a clinical data warehouse containing live Epic data from Stanford Health Care (SHC), the Stanford Children’s Hospital (SCH), the University Healthcare Alliance (UHA) and Packard Children’s Health Alliance (PCHA) clinics and other auxiliary data from Hospital applications such as radiology PACS. STARR platform is developed and operated by Stanford Medicine Research IT team and is made possible by Stanford School of Medicine Research Office. We thank Stanford Research Information Technology for their support and assistance in data acquisition and extraction. We thank the members of the Stanford Quantitative Sciences Unit for their review and feedback on the manuscript.

## Contributors’ Statements

Dr. Bannett conceptualized and designed the study, defined and coordinated data extraction, participated in chart reviews, carried out the data analyses, drafted the manuscript, and reviewed and revised the manuscript.

Dr. Dahlen participated in study design, extensively reformatted the data for analysis, performed statistical data analysis, and critically reviewed and revised the manuscript.

Dr. Huffman participated in study design and conceptualization, and critically reviewed and revised the manuscript.

Dr. Feldman supervised the conceptualization and design of the study and critically reviewed and revised the manuscript.

Drs. Bannett and Dahlen had full access to all the data in the study and take responsibility for the integrity of the data and the accuracy of the data analysis. All authors approved the final manuscript as submitted and are responsible for all aspects of the work.

